# High seroconversion rate and SARS-CoV-2 Delta neutralization in PLWHIV vaccinated with BNT162b2

**DOI:** 10.1101/2021.12.07.21267395

**Authors:** Valérie Pourcher, Lisa Belin, Cathia Soulie, Michelle Rosenzwajg, Stéphane Marot, Karine Lacombe, Nadia Valin, Gilles Pialoux, Ruxandra Calin, Isabelle Malet, Karen Zafilaza, Roland Tubiana, Marc-Antoine Valantin, David Klatzmann, Vincent Calvez, Noémie Simon-Tillaux, Anne-Geneviève Marcelin

**Affiliations:** Sorbonne Université, INSERM, Institut Pierre Louis d’Epidémiologie et de Santé Publique, AP-HP, Hôpitaux Universitaires Pitié-Salpêtrière – Charles Foix, Service de maladies infectieuses, Paris, France; INSERM, Institut Pierre Louis d’Epidémiologie et de Santé Publique, AP-HP, Hôpitaux Universitaires Pitié-Salpêtrière – Charles Foix, Département de Santé Publique, Paris, France; Sorbonne Université, INSERM, Institut Pierre Louis d’Epidémiologie et de Santé Publique, AP-HP, Hôpitaux Universitaires Pitié-Salpêtrière – Charles Foix, laboratoire de virologie, Paris, France; Sorbonne Université, INSERM Inflammation-Immunopathology-Immunotherapy Department (i3) and AP-HP, Hôpital Pitié-Salpêtrière, Clinical Investigation Center for Biotherapies (CIC-BTi), F-75013, Paris, France; Sorbonne Université, INSERM, Institut Pierre Louis d’Epidémiologie et de Santé Publique, AP-HP, Hôpital Saint-Antoinre, Service de maladies infectieuses, Paris, France; Sorbonne Université, INSERM, Institut Pierre Louis d’Epidémiologie et de Santé Publique, AP-HP, Hôpital Tenon, Service de maladies infectieuses, Paris, France; Sorbonne Université, INSERM, Institut Pierre Louis d’Epidémiologie et de Santé Publique, AP-HP, Hôpitaux Universitaires Pitié-Salpêtrière – Charles Foix, Département de Santé Publique, Paris, France

**Keywords:** COVID-19, PLWHIV, mRNA vaccine, variants, antibodies

## Abstract

**Objectives:** To assess the humoral and cellular responses against SARS-CoV-2 Delta variant after BNT162b2 vaccination in PLWHIV.

**Design:** Multicenter cohort study of PLWHIV, with a CD4 cell count <500/mm^3^ and a viral load <50 copies/ml on stable antiretroviral therapy for at least 3 months.

**Methods:** Anti-SARS-CoV-2 Receptor Binding Domain IgG antibodies (anti-RBD IgG) were quantified and their neutralization capacity was evaluated using an ELISA (GenScript) and a virus neutralization test (VNT), against historical strain, Beta and Delta variants before vaccination (day 0) and one month after a complete vaccination schedule (M1).

**Results:** 97 patients were enrolled in the study: 85 received 2 vaccine doses (11 previous COVID-19 and 1 premature exit). The seroconversion rate in anti-RBD IgG was 97% CI95[90%; 100%] at M1. Median (IQR) anti-RBD IgG titer was 0.97 (0.97-5.3) BAU/ml at D0 and 1219 (602-1929) at M1. Neutralizing antibodies (NAbs) capacity improved between D0 (15% CI95[8%; 23%]) and M1 (94% CI95[87%; 98%]) with the GenScript assay (p<0.0001). At M1, NAbs against historical strain, Beta and Delta variants were present in 82%, 77% and 84% patients respectively. The seroconversion rate and median anti-RBD IgG were 91% and 852 BAU/ml in patients with CD4<250/mm3 (n=13) and 98% and 1270 BAU/ml in patients with CD4>250/mm^3^ (n=64) (p=0.3994). 73% of patients with CD4<250 had NAbs and 97% of those with CD4>250 (p=0.0130). The NAbs against Beta variant was elicited in 50% in CD4<250 and in 81% in CD4>250 (p=0.0292). No change in CD4^+^ or CD8^+^ T cells count was observed while a decrease of CD19^+^ B cells count was observed (208 ±124 cells/mm3 at D0 vs 188 ±112 cells/mm3 at M1, p<0.01). No notable adverse effects or COVID-19 were reported.

**Conclusions:** These results show a high seroconversion rate with a Delta neutralization in PLWHIV patients after a complete BNT162b2 vaccination schedule.

## Introduction

The SARS-CoV-2 pandemic represents an extraordinary challenge for global health, it has affected hundreds of millions of people worldwide and has led to more than 5 million deaths as of december 2021. Seroprevalence of SARS-CoV-2 infection in people living with HIV (PLWHIV) is very variable depending on the country between 0.72 to 8.5% ^[1-4]^. Data on whether HIV increases the risk of severe COVID-19 are conflicting, however 3 recent meta-analyses are in favor of an associated higher risk of mortality from COVID-19 ^[5-7]^.

Vaccination against COVID-19 is an essential weapon to mitigate the consequences of the SARS-CoV-2 pandemic. Its effect in reducing a wide range of COVID-19-related outcomes including mortality, proved in the healthy population in real-word setting, is of course expected in most vulnerable people such as immunocompromised people ^[8]^.

One of vaccines authorized in France is a mRNA encoding the spike protein of SARS-CoV-2 and formulated in lipid nanoparticles called BNT162b2 (Pfizer-BioNTech). Phase 2/3 studies of this vaccine have shown its high efficacy and good tolerance ^[9, 10]^. Although this study included PLWHIV but data are lacking.

Another issue is the worldwide emergence of SARS-CoV-2 variants, since some variants partially escapes neutralizing antibodies ^[11]^, due to amino-acids deletions and substitutions in functional regions of the receptor binding domain (RBD). The Alpha (B.1.1.7) variant of concern ^[12]^ showed a 3-fold reduction of NAb titers in mRNA-vaccinated people, compared to the historical B.1 virus ^[13-15]^. The Beta VOC (B.1.351) showed a more potent neutralizing escape, with up to 14-fold decrease of NAb titers, either in vaccinated or convalescent individuals ^[13-15]^. The Delta VOC (B.1.617.2), which is now dominant worldwide showed a 3- to 5-fold decrease of NAb titers than the Alpha variant in vaccinated individuals ^[16]^. Thus, obtaining specific data on the efficacy of the BNT162b2 vaccine against Delta variant in a frail patient population such as PLWHIV is crucial.

We conducted a multicenter cohort study of people at risk of developing severe forms of COVID-19, including PLWHIV, with the main objective to evaluate the neutralizing capacity of the antibodies towards different variants of SARS-CoV-2 before BNT162b2 vaccination (day 0) and one month after a complete vaccination schedule (M1).

## Materials and Methods

### General and ethical considerations

The study follows the STROBE reporting guidelines (see checklist in supplemental table XX) ^[17]^. The study was registered on ClinicalTrial.gov (NCT04844489).

The study was approved by the ethical and scientific committee of Ile de France 8 under the registration number IDRCB 2021-A00469-32. All patients were informed and gave their written consent. As a French priority matter of research during the COVID pandemic, the study got the CAPNET label. The COVIVAC-ID study is a prospective cohort of immunocompromised patients that received the BNT162b2 vaccine, with several sub-cohorts according to the type of immunosuppression. This report is focused on patients living with HIV.

### Patients

We included PLWHIV followed in the departments of infectious diseases in the Greater Paris University Hospitals (AP-HP Sorbonne Université) with a CD4 cell count <500/mm^3^ and a viral load <50 copies/ml on stable antiretroviral therapy for at least 3 months and eligible to COVID-19 vaccination. Exclusion criteria were a previous known SARS-CoV-2 infection in the 3 last months (assessed by a positive RT-PCR, Lamp PCR or antigen test), other vaccination received in the 15 days preceding recruitment, known or suspected allergy to any component of the vaccine or severe allergic site, evocative signs of COVID-19, intercurrent infection, pregnancy in progress or expected within 3 months post-vaccination. BNT162b2 vaccine was administered according to a 2-dose vaccination schedule at an interval of 3 to 6 weeks as recommended in France at this time. Patients were followed before vaccination (day 0), after the first dose and before second vaccination (day 28) and one month after the last dose (M1). Secondary research objectives, evaluated at the same time, included evaluation of seroconversion rate, percentage of patients who had a previous infection before vaccination, evolution of the antibodies ^[10]^ level after vaccination, comparison of the neutralizing capacity of antibodies between the different variants of SARS-CoV-2, correlation between the antibodies level and the neutralization titers after vaccination, changes in T lymphocyte subpopulations and their functional markers of activation.

### SARS-CoV-2 serology, neutralizing antibodies

Qualitative detection of nucleocapsid protein (anti-N) IgG Abs and quantification of anti-spike RBD IgG Ab (SARS-CoV-2 IgG II Quant, Abbott, Rungis, France) were assessed by automated chemiluminescence assay on Abbott Alinity i platform, according to manufacturer’s instructions. An IgG index ≥ 0.8 indicates a positive serology for anti-N and the cut-off positivity for anti-S is at 7.1 binding Ab unit per milliliter (BAU/ml).

### Neutralizing Antibodies

A semi-quantitative ELISA assay (SARS-CoV-2 Surrogate Virus Neutralization Test, Genscript, USA) based on antibody mediated blockade of ACE-2-Spike protein interaction was performed as described previously ^[18]^. The more NAbs are present in the samples, the more they inhibit the binding of labeled RBD to ACE-2, which results in less signal. According to the manufacturer, an inhibition above 30% is considered positive.

The neutralizing activity of sera was also assessed by a whole virus replication neutralization test (VNT) as previously described ^[19]^ using three SARS-CoV-2 isolates: the historical (B.1), Beta (B.1.351) and Delta (B.1.617.2) strains (GenBank accession number MW322968, MW580244 and GISAID accession ID 3431813). Microscopy examination was performed on day 4 to assess the cytopathic effect (CPE). NAbs titer greater than 1:10 is considered as positive.

### Flow cytometry

Blood samples were collected in EDTA tubes according to the planned protocol: absolute numbers of lymphocyte subsets were monitored at each patient visit: day 0, day 28 and M1. Blood subsets (CD3^+^, CD4^+^, CD8^+^ T lymphocytes, CD19^+^ B lymphocytes and CD3^-^ CD56^+^ NK cells) counts (cells/μl) and percentages were established from fresh blood samples using CYTO-STAT tetraCHROME kits with Flowcount fluorescents beads and tetra CXP software with a FC500 cytometer (Beckman Coulter) according to manufacturer’s instructions.

### Data collection

All data were collected in a web-based case reported form. The following covariates were collected at inclusion : history of known COVID-19, presence of comorbidities, HIV infection duration, history of opportunistic infections, history of virologic failure, duration of HIV suppression, clinical category of HIV infection, history of illnesses associated to HIV infection, immunovirological status (VL, CD4 and CD8 counts), blood cell count, liver and kidney function tests, C-reactive protein level, and class of antiretroviral therapy. During follow-up, we also searched for any failure of COVID-19 vaccination (documented infection of SARS-CoV-2 and variant if available). All adverse events were also recorded.

### Statistical analysis

#### Sample size

The sample size of each sub-cohort of the COVIVAC-ID study is based on a hypothesis on the primary endpoint measured 28 days after the first shot. According to the published literature at the beginning of the study ^[20]^, we expected a rate of 80% of seroneutralization against the Alpha variant. Ninety-seven patients were necessary to measure this proportion with a precision (half of the 95% confidence interval) of 8% in each sub-cohort.

#### Descriptive analysis

Patients were described with quantitative variables expressed as median and interquartile range, and qualitative variables expressed as number and percentage.

#### Primary and secondary outcomes analyses

The primary endpoint was the proportion of patients with a NAb titer greater than 1:10 against the historical strain and the Beta and Delta variants, reported as number with percentage with its 95% confidence interval (Clopper-Pearson interval). The analyses were performed on the complete-cases dataset, and no imputation was performed regarding the outcomes for patients with missing data. Subgroups according to CD4 cell count at baseline was prespecified, but the categories were restricted to patients with CD4 cell count above or below 250/mm^3^, due to limited number of patients with very low values. For post-hoc analysis of patients according to their history of COVID-19, we restricted the sample of patients to those without any unknown SARS-CoV-2 infection (i.e. with anti-N antibodies at baseline, but without known clinical history of COVID-19), to compare patients with 2-shots versus 1-shot and a history of COVID-19. As serological results were not known during follow-up, patients with unknown history of COVID-19 got 2-shots of vaccines. The primary and secondary endpoints were compared between subgroups with appropriate tests according to the type of variable and their distribution, i.e. Wilcoxon rank sum tests or Student t-tests for continuous variables and chi-squared or Fisher’s exact tests for categorical variables.

All analyses were performed with the software R, version 4.0. All tests were two-sided, and the level of significance was set to .05 for each comparison. No correction for multiple comparisons was made, and the results regarding subgroups analyses remain exploratory.

## Results

### Patients

97 PLWHIV were enrolled in the study. The baseline characteristics are described in table 1. At time of vaccination, all had a VL < 50 copies/ml under ART, with a median CD4 cell count of 401/mm^3^ and an average time from HIV diagnosis of 11 years. Most of them were receiving integrase inhibitors-based therapy (71%). Among them, 85 patients received 2-shots of vaccine (11 previous COVID-19 and 1 premature exit). The median time between the 2-shots was 28 [IQR 28-29] days. 90 patients could be evaluated at M1. Tolerance was overall very good and no COVID-19 was reported until M1.

**Table 1.** Baseline characteristics of PLWHIV vaccinated with BNT162b2 vaccine (n = 97)

|  | **Median or number** | **IQR or %** |
| --- | --- | --- |
| Age, years | 48 | 39-56 |
| Sex, male | 85 | 87.6% |
| BMI | 24 | 22.0-27.1 |
| Other chronic diseases | 39 | 40.2% |
| - Obesity (BMI > 30 kg/m<sup>2</sup>) | 11 | 11.3% |
| - Arterial hypertension | 17 | 17.5% |
| - Bronchopulmonary chronic obstructive disease | 2 | 2.1% |
| - Diabete (1 and 2 types) | 2 | 2.1% |
| Allergia | 5 | 5.2% |
| Hemoglobin | 14.8 | 13.7-15.7 |
| Lymphocytes | 1592 | 1350-1870 |
| ALAT | 25 | 19-36 |
| ASAT | 24 | 19-30 |
| Duration of HIV infection, years | 11.1 | 7.6-22.2 |
| Past history of opportunistic infections | 47 | 48.5% |
| Past history of virological failure | 31 | 32.0% |
| ARV therapy duration, years | 10.4 | 6.5-17.4 |
| Duration of viral load <50 copies/ml, months | 78 | 28-123 |
| Tritherapy | 69 | 71.1% |
| Bitherapy | 22 | 22.7% |
| CD4 cell counts/mm<sup>3</sup> | 401 | 303-471 |
| Ratio CD4/CD8 | 0.7 | 0.4-0.9 |

IQR, interquartile range; BMI, body mass index; ARV, antiretroviral

### SARS-CoV-2 antibodies response

The SARS-CoV-2 antibodies response after the first and second doses are summarized in table 2. At baseline, 15/97 (15.5%) patients had positive anti-N IgG and 22/97 (23%) positive anti-RBD IgG suggesting a previous COVID-19. The seroconversion rate in anti-RBD IgG was 97% CI95% [90%; 100%] at M1. Median (IQR) anti-RBD IgG titer was 0.97 (0.97-5.3) BAU/ml at D0 and 1219 (602-1929) at M1 (Figure 1). Only 2 patients did not develop anti-RBD IgG at M1. These 2 patients were men, 56 and 59 years old, no obesity, with past history of opportunistic infections and virological failure, an undetectable plasma HIV VL for 4 months only and a low CD4/CD8 ratio (0.4 and 0.2).

**Table 2:** SARS-CoV-2 antibodies response and neutralizing activity before and after BNT162b2 vaccination.

|  | **Day 0**<br>n=97 | **Day 28**<br>n=85 | **Month 1**<br>n=90 |
| --- | --- | --- | --- |
| **Positive anti-N IgG, n (%)** | 15 (15.46%) | 10 (12.05%) | 12 (13.33%) |
| **Undetermined\* anti-N IgG, n (%)** | 6 (6.19%) | 5 (6.02%) | 4 (4.44%) |
| **Anti-N IgG Index, median (IQR)** | 0.08 [0.02-0.41] | 0.05 [0.02-0.21] | 0.07 [0.03-0.36] |
| **Positive anti-S IgG, n (%)** | 22 (22.68%) | 75 (90.36%) | 88 (97.78%) |
| **Anti-S IgG titer (BAU/mL), median (IQR)** | 0.97 [0.97-5.25] | 56.52 [20.80-138.52] | 1218.89 [602.12-1969.26] |
| **Positive neutralizing antibodies\*\*, n (%)** | 14 (14.58%) | 47 (56.63%) | 83 (94.32%) |

\* $0.50 < \text{anti-N IgG index} < 0.80$
\*\* GenScript ELISA assay
BAU; Binding Antibody Units; IQR, interquartile range

**Figure 1.**
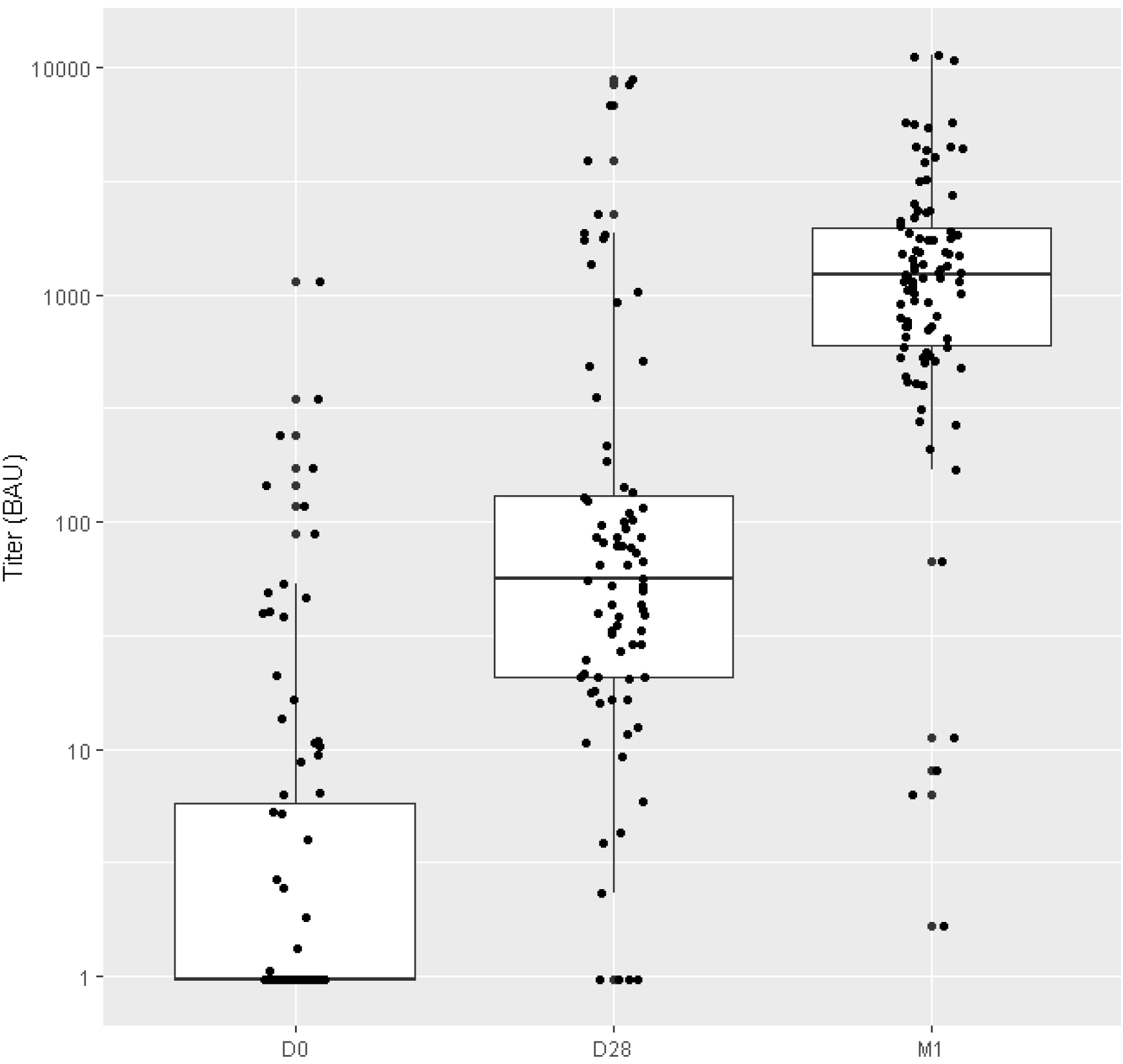
Evolution of anti-RBD IgG levels before and after first and second dose of BNT162b2 vaccine.

NAbs capacity improved between D0 (15% CI95% [8%;23%]) and M1 (94% CI95% [87%; 98%]) with the GenScript assay (p<0.0001). NAbs with the VNT were present at M1 for historical strain, Beta and Delta variants in 82%, 77% and 84% patients respectively.

Planned subgroups analysis at M1 showed that seroconversion rate and median anti-RBD IgG titer were 91% and 852 BAU/ml in patients with CD4<250/mm^3^ (n=13) and 98% and 1270 BAU/ml in patients with CD4>250/mm^3^ (n=64) (difference of change between D0 and M1 between subgroups p=0.3994). At M1, 73% of patients with CD4<250/mm^3^ had NAbs and 97% of those with CD4>250/mm^3^ (p=0.0130). There was no difference in neutralizing capacity of either historical strain or Delta variant in patients with CD4<250/mm^3^ versus patients with CD4>250/mm^3^, however the neutralizing capacity against the Beta variant was 50% in patients with CD4<250/mm^3^ and 81% in patients with CD4>250/mm^3^ (p=0.0292).

### Analysis according to previous history of COVID-19 disease

We compared the evolution of anti-RBD IgG levels and neutralization before/after vaccination according to previous COVID-19 disease or not. For this analysis, we compared patients with a known COVID-19 disease before vaccination who received 1-shot of vaccine (n=11) versus patients with 2-shots of vaccine and negative or limit anti-N and/or anti-S at D0 (n = 85). At M1, previous COVID-19 patients have a higher anti-RBD IgG level than COVID-19 naïve patients with 2345 BAU/ml [2054-4413] and 1012 BAU/ml [534-1480] respectively (p=<0.0001) (figure 2). However, both groups of patients have a high rate of neutralization at M1: 100% IC95% [76%; 100%] for patients with previous COVID-19 and 92% IC95% [83%; 97%] for naïve patients.

**Figure 2.**
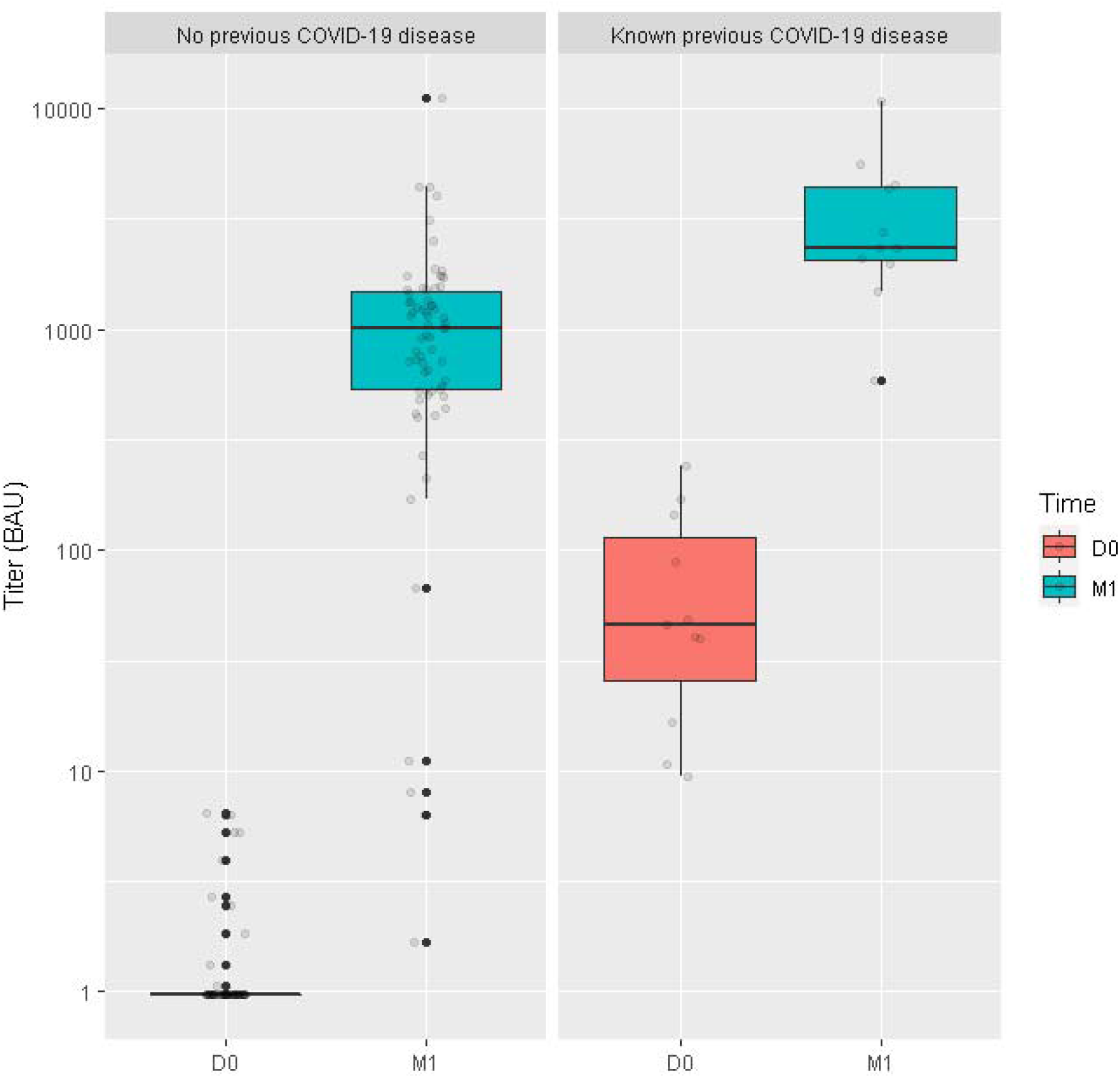
Evolution of anti-RBD IgG levels before and after the second dose of BNT162b2 vaccine according to previous COVID-19 disease or not.

### Major lymphocyte supopulation

No change in CD4^+^, CD8^+^ T, NK cells count or CD4^+^/CD8^+^ ratio was observed. Although the vaccination induced the production of antibodies, as already described in healthy people a slight but significant decrease of CD19^+^ B cells count was observed (208 ±124 cells/mm3 at D0 vs 188 ±112 cells/mm3 at M1, p<0.01) ^[21]^.

## Discussion

Our study conducted in PLWHIV with CD4 counts <500/mm^3^ and VL <50 copies/ml shows a high seroconversion rate and Delta variant neutralization elicited by a complete BNT162b2 vaccination schedule.

Several studies have been published in the past few months on the immunogenicity of COVID-19 vaccines in different immunocompromised populations. However, only few data are available for PLWHIV. Some studies conducted in PLWHIV found a low non-response rates ranging from 0 to 4% ^[11, 22]^ or comparable antibody titers compared to an HIV-negative population ^[23, 24]^. Our data confirm a prior study showing that the BNT162b2 mRNA vaccine elicits robust humoral immune responses in PLWHIV ^[24]^. We extend these data showing that in our population breadth of neutralizing antibodies, as determined by a gold-standard VNT assay with historical and VOCs, is not significantly impaired against the Delta variant.

In a recent study, mRNA-containing vaccination schemes, being female, and having a higher CD4 count was associated with a higher concentration of antibodies in PLWHIV. Particularly in the subgroup with CD4 cell counts □<□ 200/mm^3^, antibody response was poor ^[25]^. In the same way, an Italian study compared immunogenicity after a 2-shots of mRNA vaccine according to CD4 count (<200/mm^3^, 200-500/mm^3^ and > 500/mm^3^) ^[26]^. Whatever the category of CD4, the level of IgG anti-S RBD antibodies increases and is significantly different between D0 and the time of the second injection and 1 month after the second injection as in our cohort and the antibody level of all groups of HIV patients are significantly lower than the unpaired control group ^[26]^. In Yale New Haven Health System, efficacy of BNT162b2 vaccine was evaluated in PLWHIV aged ≥ 55 years at the same visits with 97.5% of PLWHIV with vaccine response with no impact of CD4 count ^[27]^. Patients enrolled in our study had CD4 counts < 500 cells/mm^3^ and we demonstrate reassuring data about vaccine efficacy in this specific population with no impact of the CD4 counts on the seroconversion rate and median anti-RBD IgG titer after a complete vaccination schedule. However, although patients with CD4 cell counts < 250 cells/mm^3^ had a retained neutralizing capacity of either historical strain or Delta variant, they had an impaired neutralizing capacity of Beta variant.

In our study, 11.3% of PLWHIV had a past history of COVID-19 infection. This in accordance with a previous study conducted in France, in May-July 2020, showing that 11% of non-HIV active workers in Paris were positive for IgG against the SARS-CoV-2 N and S proteins ^[28]^. Similarly, in 3 regions in France, including Paris, seropositivity in adults to anti-SARS-CoV-2 antibodies in May-June 2020 after the first lockdown period living, was ≤10% after the first wave and we observed the same range 10 months later in our HIV patient’s cohort ^[29]^. Comparing the antibody response after vaccination in patients with a previous COVID-19 infection who received 1-shot of vaccine and naïve patients who received 2-shots of vaccine, we found the same results than in the general population with a higher anti-RBD IgG level in previous COVID-19 PLWHIV.

Our study is limited by the relatively small number of participants and the absence of a control group. In addition, we have not yet a longer follow-up to determine the persistence of antibodies in this population. However, we assessed the neutralizing activity against different VOCs in PLWHIV.

In conclusion, we showed that PLWHIV with CD4 counts<500/mm^3^ and VL<50 copies/ml have a very good BNT162b2 vaccine response with no impact of CD4 cell count. This is highlighted by a high seroconversion rate and Delta neutralization of these patients after a complete BNT162b2 vaccination schedule.

## Data Availability

All data produced in the present study are available upon reasonable request to the authors

## Contributors

VP, LB, MR, DK, VC, NST, AGM contributed to the conception and design of the study. VP, LB, CS, MR, SM, IM, IM, KZ, DK, VC, NST, AGM contributed to the acquisition, analysis, and interpretation of data. KL, NV, GP, RC, RT, MAV enrolled patients in the study. VP, LB and AGM wrote the original draft and MR and NST substantively revised the manuscript. All authors read and approved the final manuscript.

## Acknowledgments

We thank the COVIVAC-ID staff including project manager Assitan Kone and data manager Mehdi Thamri, Sophie Sayon for her technical assistance. The sponsor was Assistance Publique – Hôpitaux de Paris (Direction de la Recherche Clinique et de l’Innovation)

